# Prediction of the effective reproduction number of COVID-19 in Greece. A machine learning approach using Google mobility data

**DOI:** 10.1101/2021.05.14.21257209

**Authors:** Arvanitis Athanasios, Furxhi Irini, Thomas Tasioulis, Karatzas Konstantinos

## Abstract

This paper demonstrates how a short-term prediction of the effective reproduction number (R_t_) of COVID-19 in regions of Greece is achieved based on online mobility data. Various machine learning methods are applied to predict R_t_ and attribute importance analysis is performed to reveal the most important variables that affect the accurate prediction of R_t_. Our results are based on an ensemble of diverse R_t_ methodologies to provide non-precautious and non-indulgent predictions. The model demonstrates robust results and the methodology overall represents a promising approach towards COVID-19 outbreak prediction. This paper can help health related authorities when deciding non-nosocomial interventions to prevent the spread of COVID-19.

## 1 Introduction

A case of unknown aetiology pneumonia was recorded in Wuhan, China, in December 2019 which escalated into an epidemic reaching a pandemic state. This novel coronavirus SARS-CoV-2 disease is named “COVID-19” by the World Health Organization (WHO). On January 30, 2020, COVID-19 outbreak was declared to constitute a Public Health Emergency of International Concern (Anastassopoulou, Russo et al. 2020). Only COVID-19 outbreak among the class of coronaviruses is declared a global pandemic in less than three months of its emergence (Salisu and Akanni 2020). The first infection in Greece was reported on the 26^th^ of February 2020, and after three days, the state applied policies to control the disease. This rapid response led Greece to be considered as a successful case of COVID-19 management compared to European and global cases (Demertzis, Tsiotas et al. 2020). For the time or writing, Greece has 281,520 confirmed cases, 8,532 deaths and 237,025 recovered cases^1^. COVID-19 infections are increasing in Greece, with 3,070 new infections reported on average each day^2^. For the time of our analysis the geographic distribution of the confirmed infected cases in Greece (145,281 confirmed cases) are shown in the map of **Figure 1** where is observed that the majority of infections are concentrated in the decentralized administration of Macedonia and Thrace.

**Figure 1.**
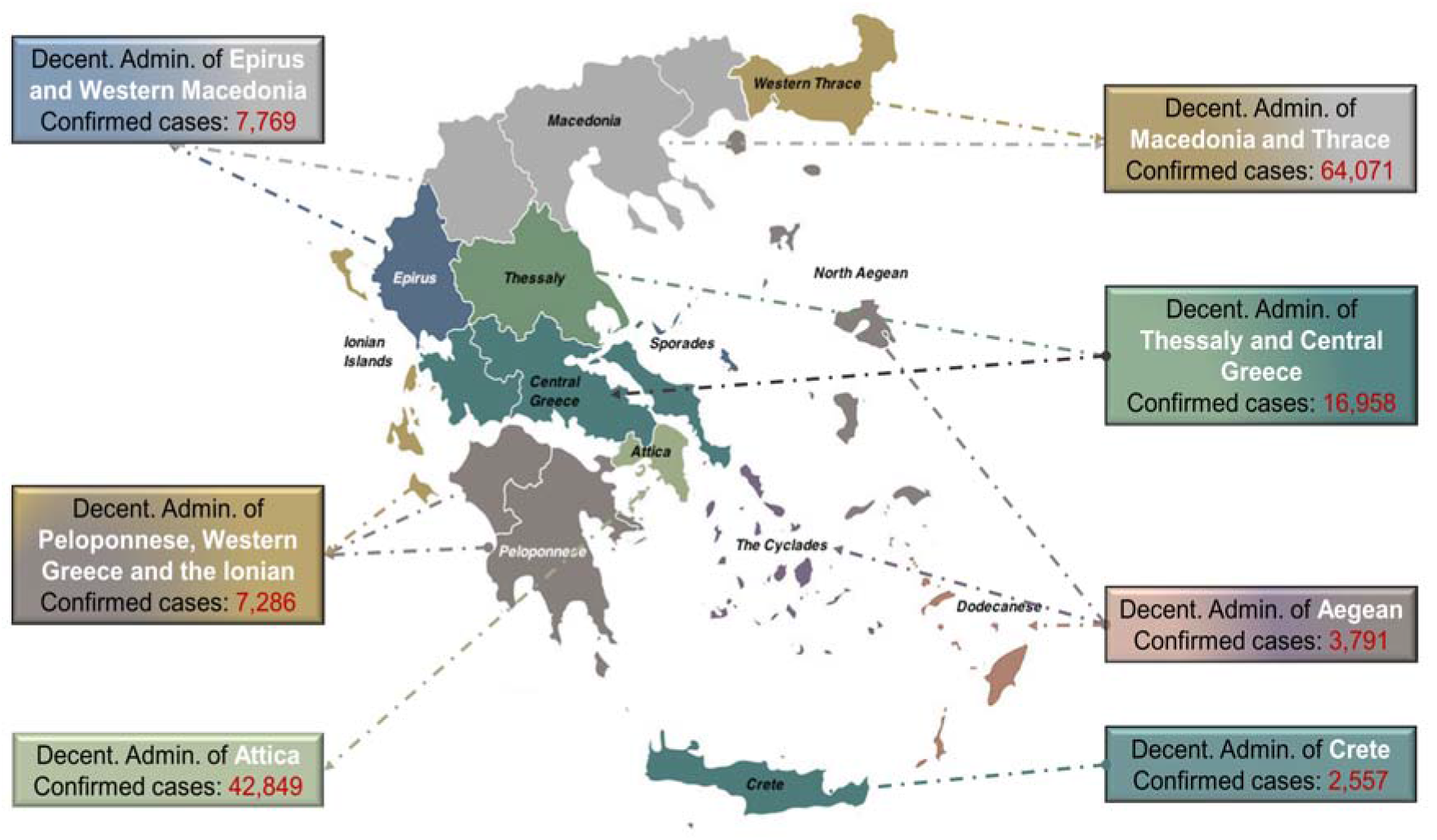
Confirmed cases in Greece across the seven decentralized administration for the period of our analysis: 18/06/2020 – 29/01/2021.

People travel and commute every day within and across countries, regions and cities. For the above reason, human mobility represents a key driver fostering the rapid spread of any disease (Wilson 1995, Tatem, Rogers et al. 2006) and has been associated with the epidemic changes of COVID-19 (Yuan, Hu et al. 2020). Consequentl, local and national governments have implemented various non-pharmaceutical interventions (NPIs) as a strategy to prevent transmission (Basellini, Alburez-Gutierrez et al. 2020, Brauner, Mindermann et al. 2020, Seale, Dyer et al. 2020).

A prominent metric that is deployed to quantify the progress of a disease is the reproduction number R (expected number of secondary cases caused by a primary case). The beginning of the epidemics is characterised by the *basic reproduction number*, R_0_ that shows the average number of secondary cases produced by a primary case when population is fully *susceptible*. Susceptible refers to individuals who can get infected if exposed and become hosts (Binti Hamzah FA, Lau C et al. 2020). Whereas, the *effective reproduction number*, R_t_ describes its progression in time. The R_t_ varies over time due to the depletion of susceptible individuals as well as changes in other factors, including mobility patterns, social mixing, control measures, contact rates, immunity of the population and climatic conditions (Arroyo-Marioli, Bullano et al. 2021). R_t_ exhibits complexity, but it is broadly accepted that values > 1.0 indicate a rapid expansion of infections (Karnakov, Arampatzis et al. 2020).

Different approaches have been proposed to model the spread of infectious diseases, such as established epidemiological models e.g., Susceptible, Infectious, Recovered (SIR) model and its extensions: Susceptible, Infectious, Recovered, Deceased (SIDR) and Susceptible, Exposed, Infectious, Recovered (SEIR) models (Ahmad, Garhwal et al. 2020). The above tools are generally based on differential equations and are still used as the baseline for comparing other approaches to epidemic modelling (Georgiou 2020). A more recent approach is based on Machine Learning (ML) tools. Diverse algorithms have been exploited such as Random Forest (RF), k-Nearest Neighbours (kNN), Support Vector Machine (SVM), Neural Networks (NN), Decision Trees (DTs), Auto-Regressive Integrated Moving Average (ARIMA) etc., Those tools have been applied to predict various COVID-19 related outcomes, for instance the number of confirmed positive cases (Saba, Abunadi et al. 2021), diagnosis and prognosis, patient outcome prediction, tracking and predicting the outbreak, drug development, vaccine discovery, false news prediction, etc (Ahmad, Garhwal et al. 2020). In addition, agent-based modelling approaches have been suggested to analyse measures and policies that are most appropriate for COVID-19 disease management for large populations (Tuomisto, Yrjölä et al. 2020)

Various computational models and approaches have been used for the case of Greece solely. Georgiou (2020) developed data analytics procedures, spectral decomposition and curve-fitting formulations. The standard epidemic modelling provided hints for the outbreak progression. Politis and Hadjileontiadis (2020) used a combination of Monte Carlo simulations, wavelet analysis and least squares optimization to a basis of SEIR compartmental models. The authors resulted in the development of stochastic epidemiological models calibrated with the epidemiological data., capable of estimating parameters of importance such as the reproduction number and the magnitude of infection. Rachaniotis, Dasaklis et al. (2021) proposed a two-phase stochastic dynamic network compartmental model (a pre-vaccination and a post-vaccination SVEIR) to assess scenarios of different phases of lockdown coupled with different vaccination rates.

Ensemble of methodologies have also been showed. Demertzis, Tsiotas et al. (2020) proposed a hybrid method based on a conceptualization of detecting connective communities in Greece in a time-series and a spline regression model where the knot vector is determined by the community detection in the complex network. Katris (2021) generated a time-series based statistical data-driven procedure to track an outbreak in Greece. The time series models include Exponential Smoothing and ARIMA approaches as classical models, also Feed-Forward Artificial Neural Networks and Multivariate Adaptive Regression Splines as ML approaches.

The R_t_ has been used in modelling approaches since it has been showed that during the evolution of the COVID-19 outbreak (in Italy), when various interventions were in place, R_t_ had a clear relation with the explosive timescale characterizing the dynamics of the outbreak (Patsatzis 2021). Linka, Peirlinck et al. (2020) using the R_t_ of COVID-19 across Europe, explored the effectiveness of political interventions proposing a dynamic SEIR epidemiology model with a time-varying R_t_ identified by ML. Kaloudis, Kevrekidis et al. (2020) provide estimations of R_t_ for Athens during the first wave of the pandemic using data from the national database of SARSCoV2 infections in Greece, by implementing commonly used methods for the estimation of R_t_, that by (Cori, Ferguson et al. 2013) and (Wallinga and Teunis 2004).

To estimate R_t_ of an infectious disease, several alternative approaches have been suggested in the literature. An extended method that of (Cori, Ferguson et al. 2013), to estimate R_t_ and its application to routine surveillance data from Greece is demonstrated by (Lytras, Sypsa et al. 2020). The resulting R_t_ estimates were examined in relation to control measures, in order to assess their effectiveness and were associated as a measure of transmissibility to population mobility as recorded in Google data and to ambient temperature. The authors conclude that mobility patterns significantly affect R_t_. Arroyo-Marioli, Bullano et al. (2021) exploited a structural mapping between R_t_ and the growth rate of the number of infected individuals derived from a SIR model while also using mobility data from Google. To compute Rt, a method proposed by (Salas 2021) starts with a form of the renewal equation of the birth process especially suitable to compute R_t_. After showing that one can express it as a linear system, they proceed to solve it, along with appropriate constraints, using convex optimization.

In this study we conduct an exhaustive application of methods to estimate the R_t_ for Greece. The R_t_ is derived from confirmed positive cases extracted from verified sources. After the estimation of R_t_, we couple the data with publicly available geo-located smartphone data provided by Google COVID-19 Mobility Reports (GCMR). This data can be used to compare pre-pandemic (baseline) to activity throughout the pandemic to better understand mobility patterns by inferring the location visited by individuals. Google community mobility data have been previously used in many other studies (Bryant and Elofsson 2020, Huynh 2020, Ilin, Annan-Phan et al. 2020, Sulyok and Walker 2020, Tamagusko and Ferreira 2020) and highlighted as one of the best data sources to analyse mobility patterns (Drake, Docherty et al. 2020). The data are used to train diverse algorithms to predict R_t_. Prediction of R_t_ is essential for public policy decisions during a pandemic. Such estimates can be used to study the effectiveness of non-pharmaceutical interventions (NPIs) or assess what fraction of the population needs to be vaccinated to reach herd immunity. Some social scientists have argued that R_t_ < 1 should be viewed as a fundamental constraint on public policy during the current COVID-19 pandemic (Arroyo-Marioli, Bullano et al. 2021).

## 2 Materials and methods

In this section, we describe the workflow for the model implementation (**Figure 2**). The first step is the data collection/extraction (***section 2.1***) from the Greek Ministry of Health, regarding the number of positive cases, and Google COVID-19 data site for the mobility change data. Once the number of cases is extracted for the geographical and temporal resolution needed, the effective reproduction rate, R_t,_ is estimated according to various methods (***section 2.2***). Data pre-processing including data gap filling by regression, data transformation, normalization and dataset random splitting to prepare the input data for the regression models (***section 2.3***). Finally, diverse ML algorithms are explored, validated and compared. Attribute importance analysis is performed to reveal the most important factors in predicting R_t_ for the best performing model (***section 2.4***).

**Figure 2.**
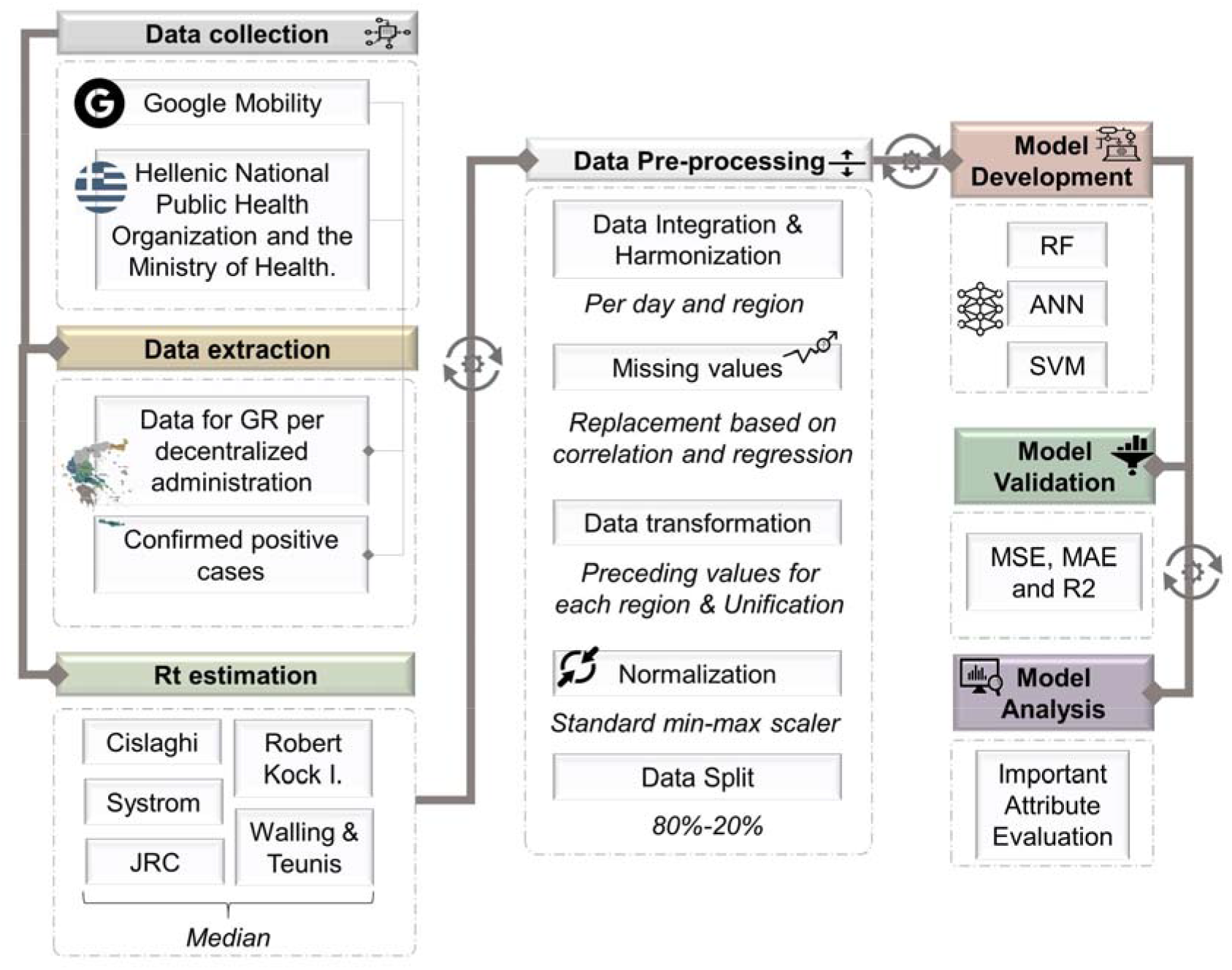
Workflow for model implementation.

### 2.1 Data Collection

#### 2.1.1 Google Mobility

Google released a time-limited sharing of mobility data from across the world as represented by summary statistics to combat COVID-19. The freely-available Google COVID-19 Community Mobility Reports (GCMR) were extracted which offers an approximation to the changes in mobility due to different social distancing measures. The GCMR accounts for the percentage changes in mobility of Google Maps users compared to a baseline period before the pandemic (from January 3 to February 6, 2020) in various categories. The movement change trend include six categories: *grocery and pharmacy, parks, retail and recreation, transit stations, workplaces* and *residential*. Each high-level category contains many types of places, for example *parks* category include information derived from public gardens, campgrounds, castles, national forests etc., or *transit stations* include seaports, subway stations, taxi stands etc.

By calculating a set of seven baseline weekdays using the median value for each individual weekday during the 5-week baseline period, the data accounts for weekly movement seasonality. For any given data date, daily relative change is valued as the percentage change with respect to the corresponding baseline weekday. GCMR for Greece provide county and decentralized administration level information. Greece is divided into seven decentralized administrations: of Attica, or Macedonia and Thrace, of Epirus and Western Macedonia, of Thessaly and Central Greece, of Peloponnese, Western Greece and the Ionian, of the Aegean and of Crete (**Figure 1**). These Google reports are created with aggregated, anonymized sets of data from users who have turned on the Location History setting, which is off by default.

Google data curator advice to not compare mobility data from different geographical regions limited this analysis to one country only. The size, economic, administrative and cultural homogeneity of Greece suggests using the mobility data for all its administrative units in a merged dataset.

#### 2.1.2 COVID-19 cases

COVID-19 data on the daily number of new reported confirmed cases were extracted from iMEdD lab GitHub^3^ as part of the development of iMEdD Lab’s web application that tracks the spread of COVID-19 in Greece and around the world. The data are sourced from verified sources such as Hellenic National Public Health Organization (EODY) and the Ministry of Health. All cases are laboratory-confirmed with RT-PCR testing, and were considered imported if they had arrived in Greece in the last 3 days before a swab was obtained (Lytras, Sypsa et al. 2020). The data included in GitHub have been originally released through press briefings announcements and transcripts published by EODY and the Ministry of Health, relevant reports published by EODY and the daily press announcements about the geographical distribution of the cases reported daily, since mid-June, 2020.

Due to the low number of COVID-19 tests, the insufficient nationwide coverage of case monitoring, the uncertainty of initial tests and irregularities in case reporting in the beginning of the pandemic, we used data from mid-June 2020 to the end of January 2021^4^.

### 2.2 Estimation of Rt

The basic reproduction number R_0_ is the number of secondary cases which one case produces in a completely susceptible population. It depends on the duration of the infectious period, the probability of infecting a susceptible individual during one contact, and the number of new susceptible individuals contacted per unit of time (Dietz 1993). It is of practical importance to evaluate time-dependent variations in the transmission potential of infectious diseases. The time course of an epidemic can be depicted by estimating the effective reproduction number, R_t_, defined as the apparent average number of secondary cases per primary case at calendar time t (for t >0). R_t_ varies in time as susceptible individuals number decreases (intrinsic factors) and control measures are implanted (extrinsic factors). If R_t_ <1, the epidemic is in decline and may be regarded as being under control (Nishiura and Chowell 2009).

There are a number of methods for estimating Rt from registered positive cases. Under the EC JRC collaboration with ECDC to continuously maintain a map18 of the level of COVID-19 transmission at sub-national level (EU 2020, EU 2020a), the Science Hub gathered and compared the most imperative methodologies of R_t_ estimation. We base our R_t_ calculations on that methods and software compilation *(JRC 2020)*. The six methods used in the JRC Technical Note are:

- Method 1: The Systrom Method. Kevin Systrom based his method on the works of (Bettencourt and Ribeiro 2008). The original method is an extension of standard epidemiological models for emerging infectious diseases, that describes the probabilistic progression of case numbers due to both human transmission and multiple introductions from a reservoir. Systrom modified the method using Bayes statistics to adjust expectation of R_t_ from the new information of each day’s case count while limiting accounting of previous data to the last seven days.
- Method 2: The Cislaghi method. The Cislaghi method uses a simple estimation of R_t_ based on new and previous case numbers applying moving averages instead of actual values (Giraudo, Falcone et al. 2020). The aim of averaging is to account for the lag between infection time and positive diagnosis time since the data available refer to the latter. We calculate R_t_ using moving averages center to 3, 4, 5 and 6 days; values between the 4 and 5-day estimates correspond to a 4.5 day incubation period^5^.
- Method 3: The JRC method. The JRC estimation is based on the basic reproduction number definition equations and the differences of the logarithms of the positive cases numbers in the formulas (JRC 2020). The method assumes a generation time of 7-days and susceptibility for the whole population.
- Method 4: Robert Koch Institute (RKI) method. The RKI method assumes a constant generation time of four days. The R_t_ is calculated as the sum of new reported cases during four consecutive days divided by the sum of new reported cases during four consecutive days prior to the last four days, using a total of eight days for determining Rt^6^
- Method 5: Exponential growth / Wallinga & Lipsitch method. The Wallinga & Lipsitch method estimates R_t_ using the exponential epidemic growth rate and the generation interval distribution. The method assumes that the growth rate follows a random distribution (Wallinga and Lipsitch 2007). Alternatively, a simpler variation of exponential growth was used with a constant rate and a time interval of one week.
- Method 6: Wallinga & Teunis method: In the Wallinga & Teunnis method the relative likelihood of a case being infected by another case is estimated assuming a probability distribution of the generation interval (Wallinga and Teunis 2004). All likelihoods summed up provide R_t_. This method failed to produce results in our case since it was impossible to fit a probability distribution to the subnational data due to discontinuities in case reporting in the less populated regions.

In total eleven estimates of R_t_ are generated using the JRC, RKI, Exponential growth-constant methods, the Systrom method with its upper and lower estimates, the Walling & Lipsitch method and the Cislaghi method for 3, 4, 5 and 6-day incubation period. In order to use one value for the prediction model we used the median of the eleven estimates.

The estimation of R_t_ was based on the programs developed by the JRC^7^ in R, modified accordingly to account for subnational resolution, i.e., the administrative units, and to extract the relevant data. The same parameterisations were used for implementing the models as in the JRC report, e.g., generation time when constant was set to 7 days and where a random distribution could be used, gamma distribution with a mean of 6.6 days and a variance of 1.5 was used.

### 2.3 Data pre-processing and Analysis

#### 2.3.1 Integration & harmonization

For this research, an integration of GCMR and COVID-19 confirmed cases is performed to acquire daily region specific information. The GCMR *Residential* category shows a change in duration (time users spent at home, using the home addresses provided to or estimated by Google Maps) —the other categories measure a change in total visitors (Lapatinas 2020). As people spend most of the day at places of residence, the ability for variation is not significant^8^. The residential data were excluded from the GCMR as expressing a different metric and due to its strong dependence on other variables used in the Google dataset. The resulting five movement change categories are *grocery and pharmacy, parks, retail and recreation, transit stations and workplaces*.

#### 2.3.2 Missing values

The original dataset was consisted of 1557 rows from which 133 had at least one missing value to one of the mobility categories. A multivariable linear regression between selected features was performed, based on existing correlation, to impute missing values.

The correlation between the mobility features in the dataset are shown in **Figure 3**. It is evident that *Park* manifest the highest correlation with *Retail & Recreation* and *Transit* features (coefficient of determination 0.94 and 0.87, respectively) and vice versa. Based on this finding, *Transit* and *Retail & Recreation* were selected for the estimation of *Park* and *Work* missing values.

**Figure 3.**
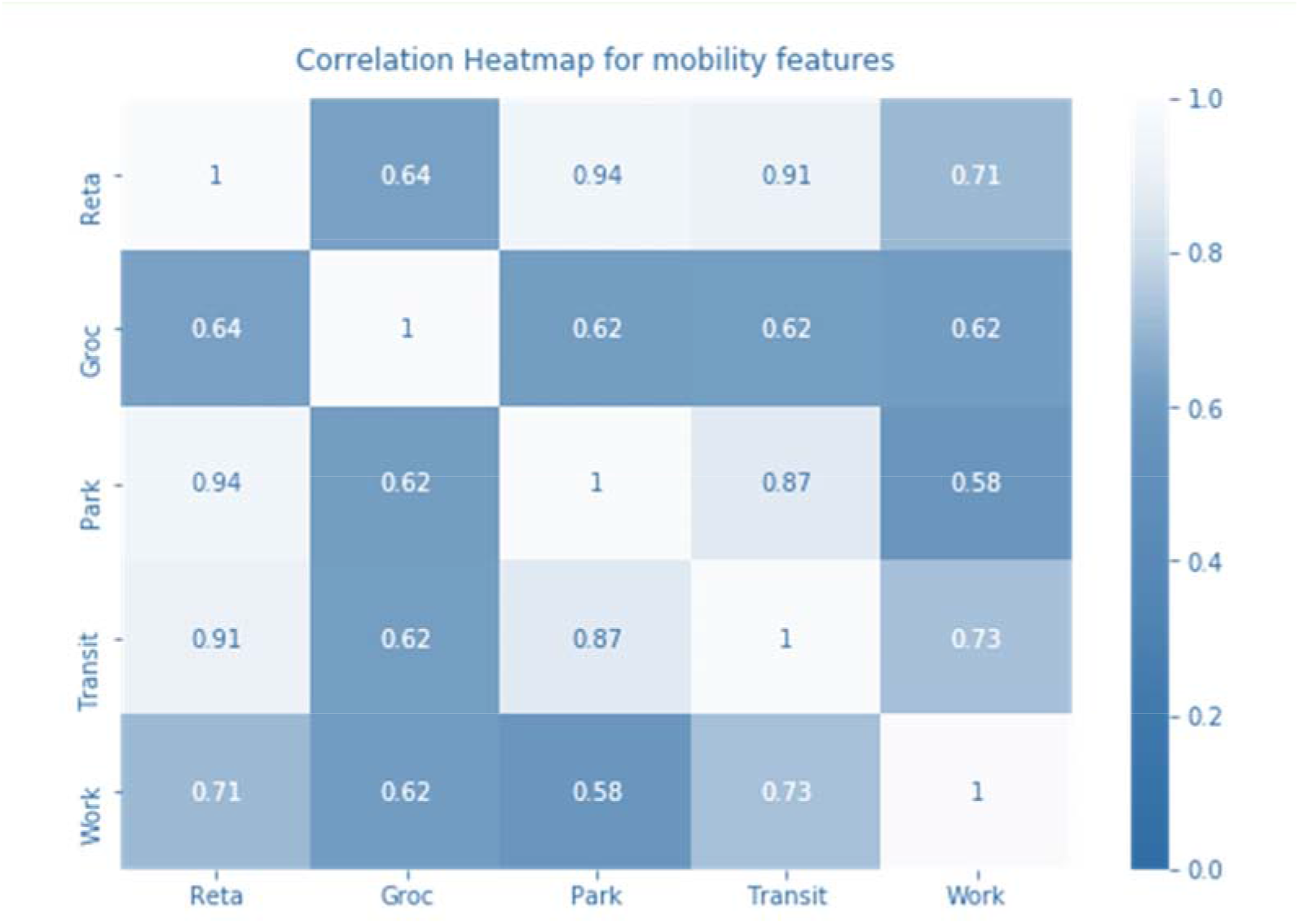
Correlation Heatmap of Mobility Features.

The estimation was performed regardless of region. Features for which the correlation was not high enough to underpin an estimation remained as such and the missing values rows were excluded from the dataset. After filling in missing values with fitted results, 62 rows remained with at least one NA value.

#### 2.3.3 Data transformation

##### 2.3.3.1 Input days

We chose the R_t_ of a day as the desired prediction outcome from the mobility data of the days of prior week. We extracted the preceding values for each day of the week for each activity and therefore, 35 features (inputs) were created to be implemented to the models. i.e. 5 features for each one of the preceding 7 days of the week: ., each input row includes preceding Monday values (*Park, Grocery, …, Workplaces)*, preceding Tuesday values, etc, up to preceding Sunday values. Since the mobility data are defined as change based on reference values per day, a certain percentage change on a Monday can only be compared to another Monday percentage. For instance, the high percentage change in Grocery-Sundays reflects that more grocery shops were open on Sundays during the pandemic, but those peak values should not be combined with Saturday mild changes in one model attribute, since they reflect different social behaviours. For this reason, instead of grouping the mobility data to input attributes according to whether they refer to one day, two days, etc, before current day, we grouped them in preceding Monday, preceding Tuesday, etc. Current day measurements of mobility were excluded from the input attributes to be able to investigate if R_t_ can be prognosed (forested), not just estimated (nowcasted).

#### 2.3.4 Normalization

Data values were normalized to help the training process using a min-max scaler where the highest value corresponds to 1 and the lowest value to 0.

#### 2.3.5 Data Split

The dataset was split into train and test sub-sets, at 80% and 20% for training and external validation, respectively. To make one-day predictions, 1288 rows were used which were randomly split to 1030 and 258 rows for train and test, respectively. In Artificial Neural Network (ANN) model, the train set was further split randomly to train and validation set with a proportion of 80 to 20; (not including here the intrinsic cross-validation splitting).

### 2.4 Modelling approach

#### 2.4.1 Machine learning tools

Three different machine learning (ML) algorithms were trained to predict R_t_ for each day, Random Forest (RF), Artificial Neural Network (ANN) and Support Vector Machine (SVM). Models are built in Python version 3.7.6, Scikit-learn version 0.24.1 and TensorFlow 2.1.0 (for ANN).

Numerous studies have used RF for either classification or regression across various domains (Mutanga, Adam et al. 2012, Smith, Ganesh et al. 2013, Sarica, Cerasa et al. 2017). The algorithm has been successfully applied in modelling studies concerning COVID-19 spreading (Singh, Kumar et al. 2020). RF is an ensemble-based meta-classifier employing decision trees, used here for regression purposes (Breiman 2001). RF randomly chooses features and applies resampling throughout the time stamped records with replacement (bootstrapping). On this basis, a regression tree is trained for each subset and the average of their predictions is the output of the model. The main advantage of RF is the measurements of importance for each *k* predictor which although similar to multivariate linear regression it outperforms the latter in almost all non-linear applications (Grömping 2009). A total of 10,000 trees was used in this study. The maximum pruning depth was left without control in order to keep individual error low, according to (Segal 2004). The forest was built using bootstrap aggregation which is primarily used for unknown distributions and constitutes a standard basis to reduce variance and bias on a sample (Efron 1979). The cost function was chosen as Mean Squared Error (MSE) because, after trial, it showed significantly better results during the training process.

ANN is an universal function approximator that has been widely used in various epidemiological and environmental studies (Philemon, Ismail et al. 2019). In this study, ANN was built using 3 layers without initial bias. The first layer was consisted of 250 nodes, the second of 125 nodes and the third of 60 nodes. All three layers used rectified linear function, whereas on the first and third layer we applied a 10% dropout of training samples to avoid overfitting (Srivastava, Hinton et al. 2014). The 3-layer architecture was chosen to reduce the bias of the model caused by collinearity within the features. The number of nodes for each layer was estimated by multiple trials with a principle of narrowing down the nodes after each layer; this choice of less nodes at each layer enabled the model to evaluate higher level features and reduce bias, achieving better predictions of R_t._ MSE was chosen as the loss function. The learning rate was set to 0.001 and *Adam* optimizer was implemented, as it is widely used on noisy data (Kingma and Ba 2017) and outperforms stochastic gradient descent during training.

SVMs are based on the principle of data separation via a hyperplane that optimises data division, and has been suggested for COVID-19 modelling purposes (Gupta, Singh et al. 2021). The regression tasks is made by estimating data points close to the hyperplane (support vectors) and minimizing the distance between those datapoints to a selected threshold epsilon (*E*), corresponding to error of tolerance. With regard to hyperparameters, regularization factor (C) was set to 100 in order to reduce margin boundaries and achieve better accuracy rate in terms MSE (Chang, Hsieh et al. 2010). The best kernel was found to be the Radial Basic Function (RBF) kernel, compared to the polynomial and linear kernel (data not shown). *E* was set to 0.2 corresponding to the lowest amount of MSE of tolerance.

#### 2.4.2 Validation

A 10-fold cross-validation was used during the internal training (10 times random 90%-10% splitting for training and testing); the external evaluation of the model was performed with a test set (random 20% of the whole dataset consisting of data points not implemented in the training). Various metrics were calculated such as the MSE, Mean Absolute Error (MAE), R squared (R^2^), Pearson’s and Spearman’s correlation (Kuo and Fu 2021).

#### 2.4.3 Important Attribute Analysis

Attribute importance was derived through random forest optimization (built-in function) to reveal the most significant variables that relatively affect the prediction of the outcome. The analysis is based on the Gini importance, an all-nodes accumulating quantity that indicates how often a particular attribute was selected for a split, and how large its overall discriminative effect was in the regression (Menze, Kelm et al. 2009).

## 3 Results

### 3.1 Estimation of Rt

**Figure 4** demonstrates an example of the R_t_ estimations of the various methods employed in this study and the estimated median, for the administration unit of Macedonia and Thrace.

**Figure 4.**
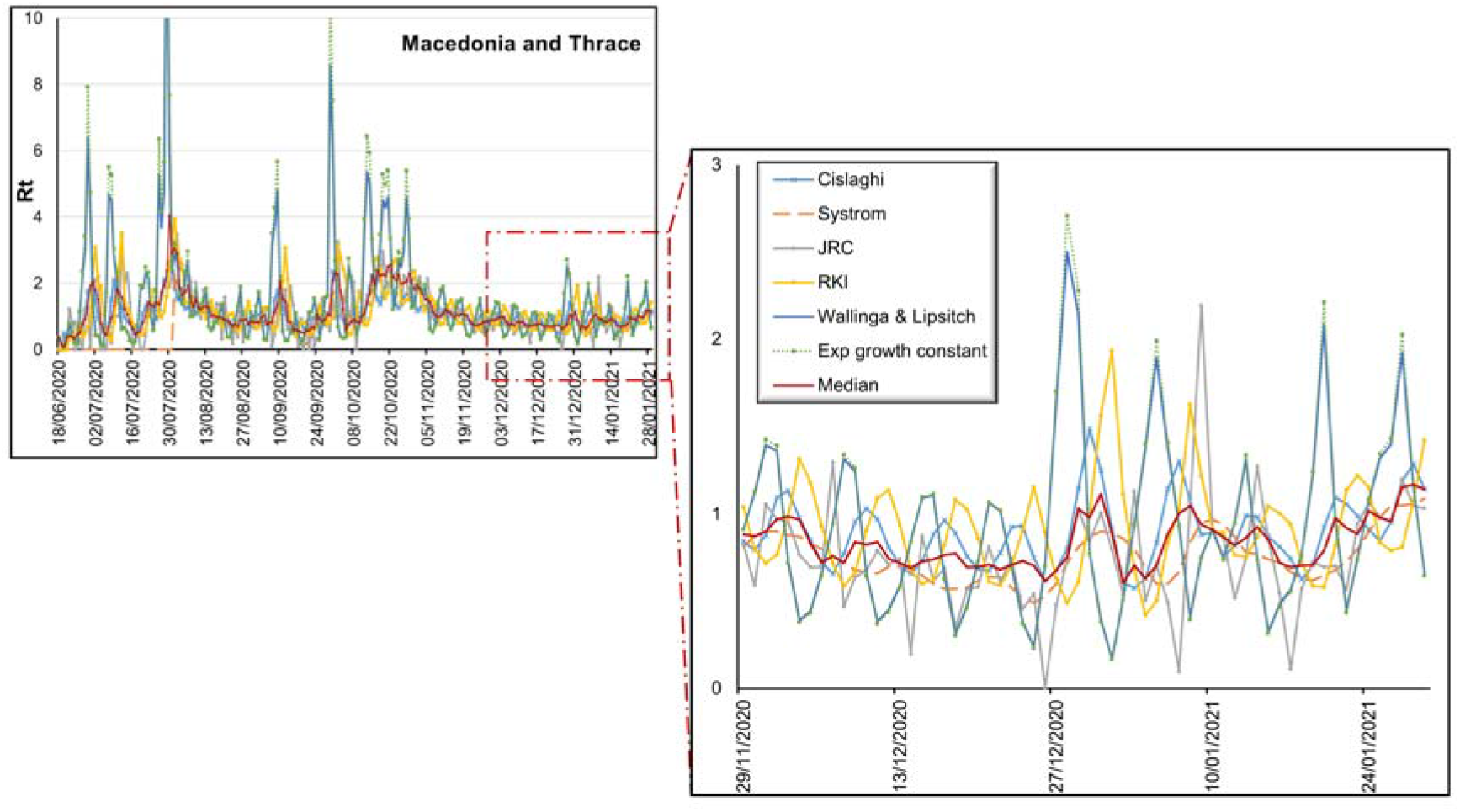
Rt estimations of the Macedonia and Thrace region using various methods. The zoomed picture facilitates the visual inner-comparison.

*Cislaghi* (light blue), *Systrom* (orange) and *JRC* methods (grey) are close to their estimations affecting the most the median line (red) that shows similar variance and concurrent peaks. JRC is generally a little bit below the other methods, while Cislaghi is always a bit higher. Systrom is the smoothest and with lower average estimates.

*Wallinga & Lipsitch* (dark blue) and *Exp growth constant* (dotted green) R_t_ are almost identical representing same trends and reaching the highest, unrealistic peaks (>10). Contrary to the others, those two methods give R_t_ above 2 for the Fall pandemic wave. They demonstrate the greatest divergence of values although *Wallinga & Lipsitch* cuts off maximum values compared to the constant growth rate exponential method.

*Systrom* (orange) is smoother with lower average estimates. On the other hand, *RKI* (yellow line) shows peaks shifted in time compared to other methods, which are sharp and closer to the exponential methods but with lower variance.

Due to the time shifts on the peaks the median line (red) smooths down the estimates and provides a more realistic and continuous evolution.

### 3.2 Modelling approach and validation

The results of RF, ANN and SVM model performance metrics for the test subset are shown in **Figure 5**. RF demonstrates the lowest MSE and MAE in comparison to SVM and ANN. It also achieves the highest R^2^, Pearson’s and Spearman’s correlation values, outperforming in all metrics the other models. RF has been demonstrated to outperform other basic classifiers, even with missing values present in the dataset (Furxhi, Murphy et al. 2019, Furxhi and Murphy 2020).

**Figure 5.**
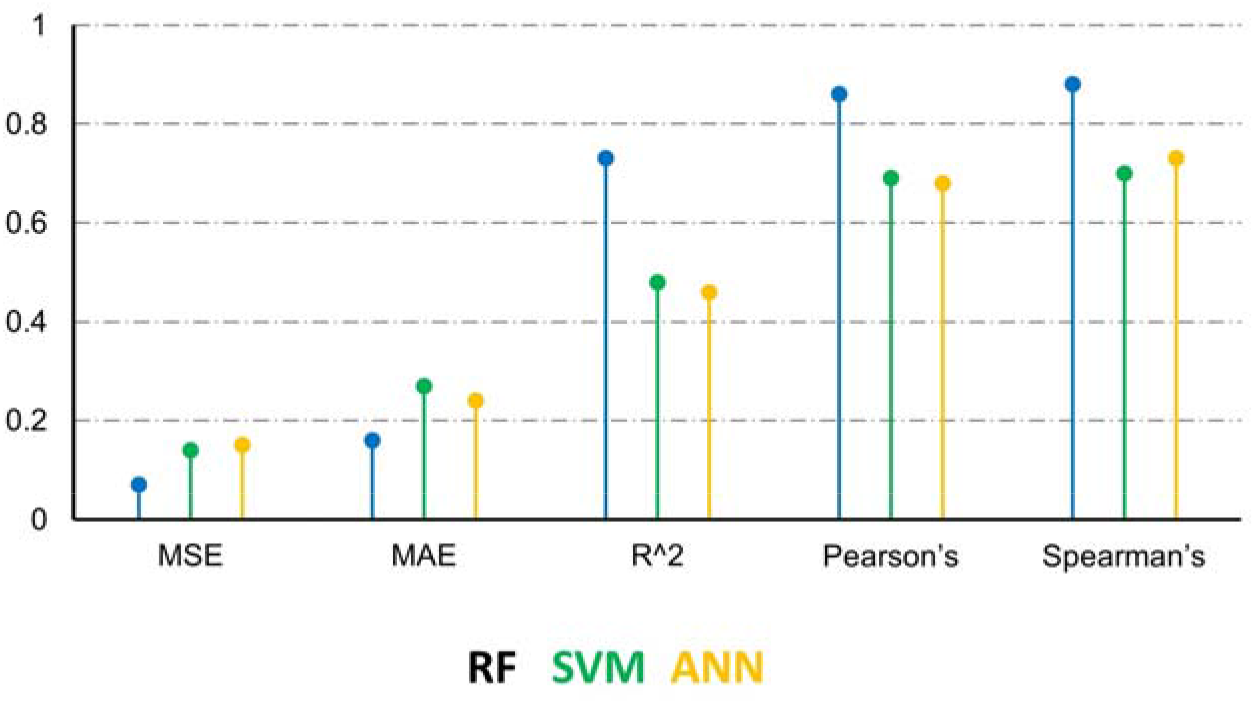
Validation metrics of the RF(blue line), ANN and SVM (green line) and ANN (yellow line) models.

SVM and ANN have similar performance with R^2^ below 0.5 and comparable Pearson’s and Spearman’s around 0.7. RF achieves MAE below 0.2 and MSE below 0.1, maintaining Spearman’s and Pearson’s close to 0.9.

R^2^ measures the variance of the model. RF is able to generalize the predictions regardless of the peak values and can isolate outliers more efficiently mainly due to the inherent ensembling nature of the algorithm. On the other hand, ANN and SVR achieve decent Pearson’s and Spearman’s coefficients (i.e., above 0.6), but are affected more by outliers, peak values and high variance in the features. Spearman’s correlation is slightly higher than Pearson’s for every model showing that each model is able to capture the trendline of R_t_ scoring only marginally lower in terms of variance.

The predicted R_t_ values are plotted in **Figure 6**. It is evident that RF predictions are less scattered compared to the observed^9^ data. In case of low R_t_ values (<1.0), RF is precautious and predicts slightly higher values. The opposite trend is noticed when the observed values are higher than 1.0. SVM and ANN show a similar pattern. SVM underestimates above 1.3 and ANN above 0.8. However, both models show more scattered predictions.

**Figure 6.**
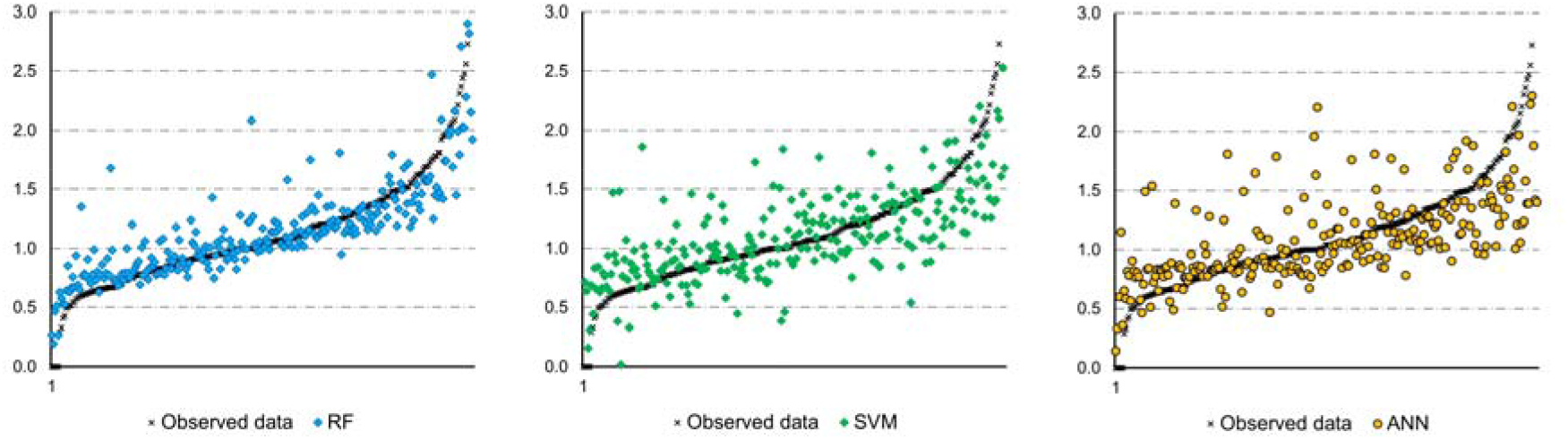
RF, SVM and ANN external predictivity.

**Figure 7-Figure 13** show ordered by time and geographic location how model results compare to observed R_t_ for the test dataset (which has been selected by random sampling according to chapter **2.4.2** and therefore containing non sequential dates).

**Figure 7.**
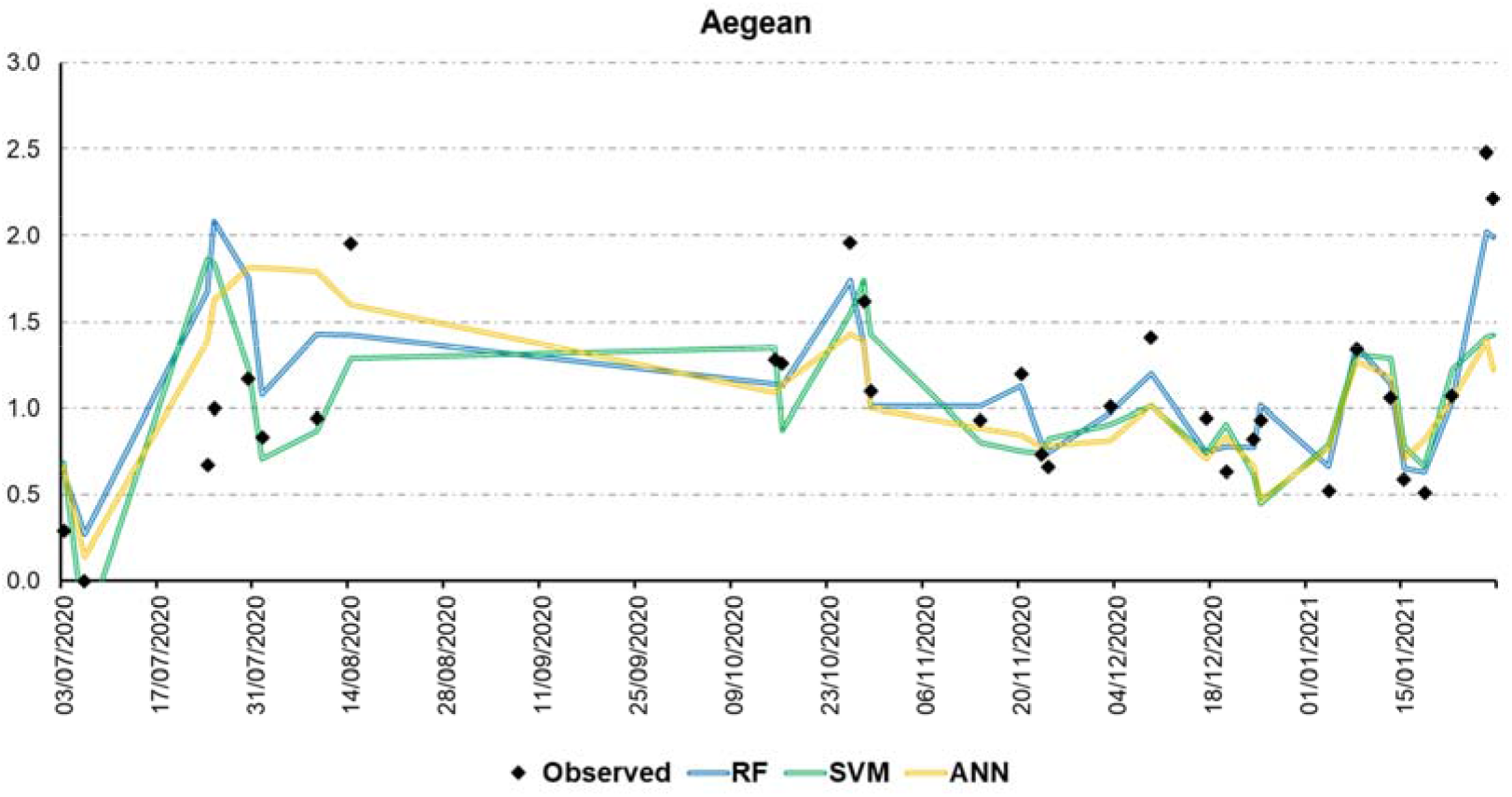
Predicted vs Observed Rt for the Aegean region.

**Figure 8.**
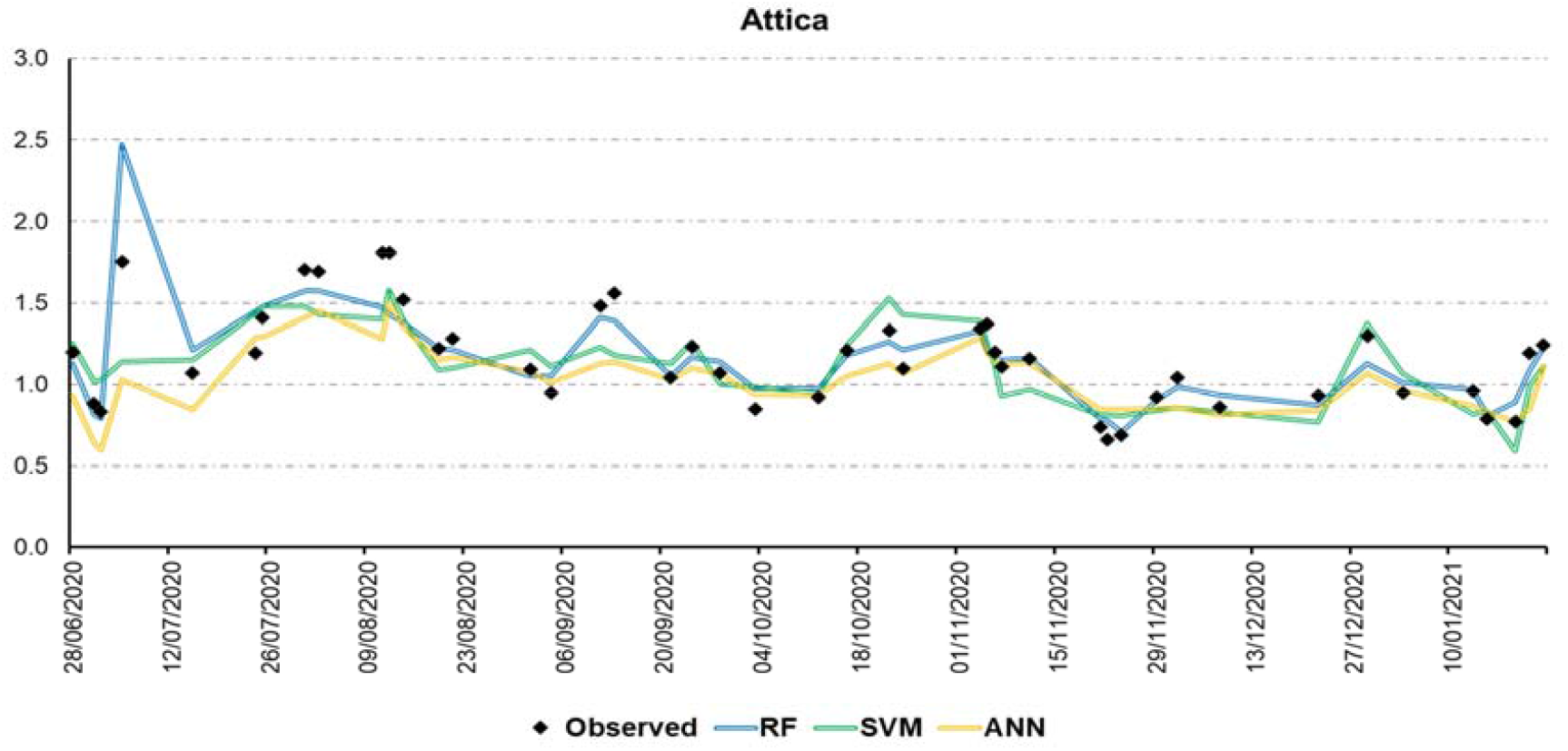
Predicted vs Observed Rt for the Attica region.

**Figure 9.**
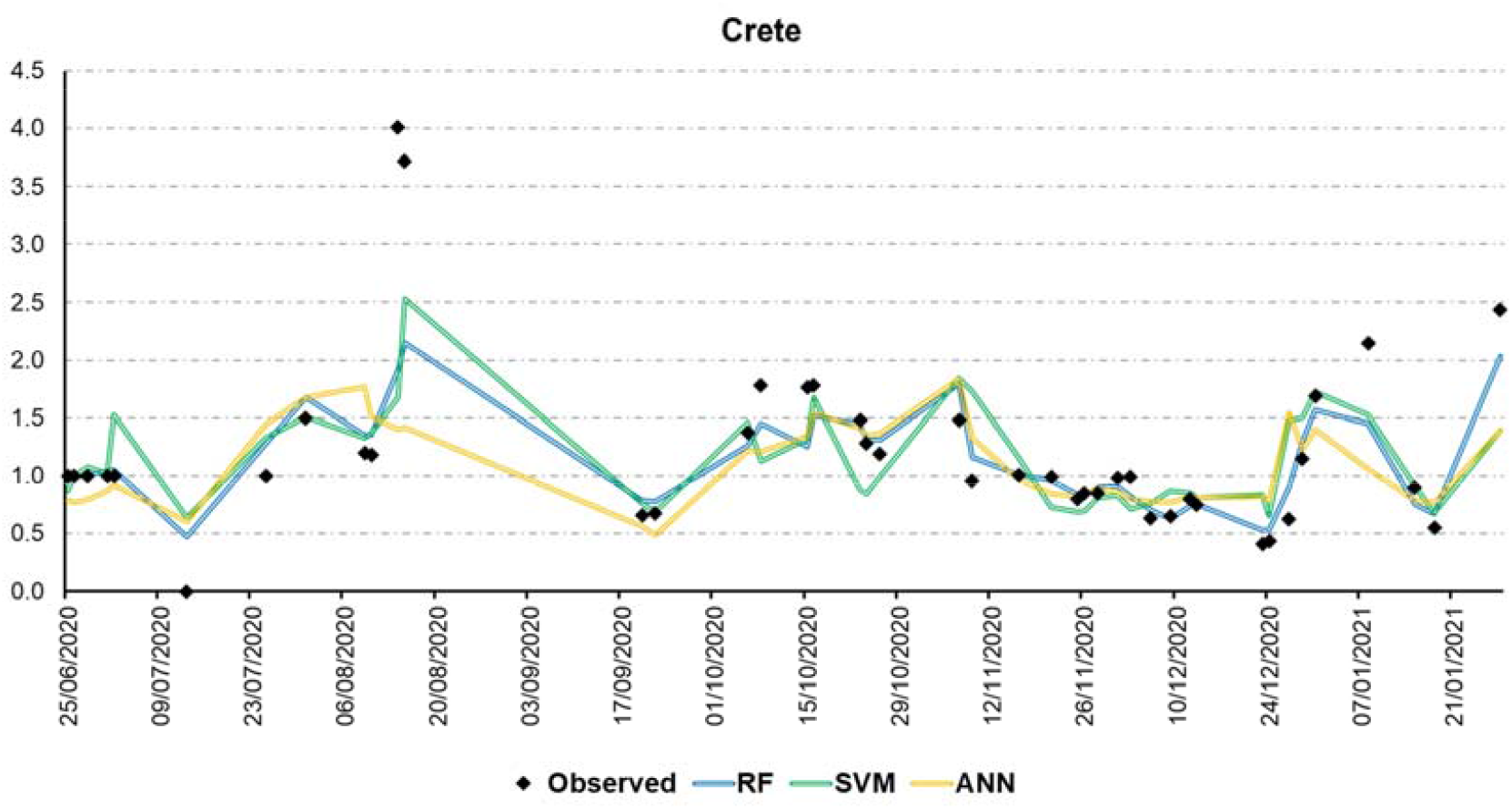
Predicted vs Observed Rt for the Crete region.

**Figure 10.**
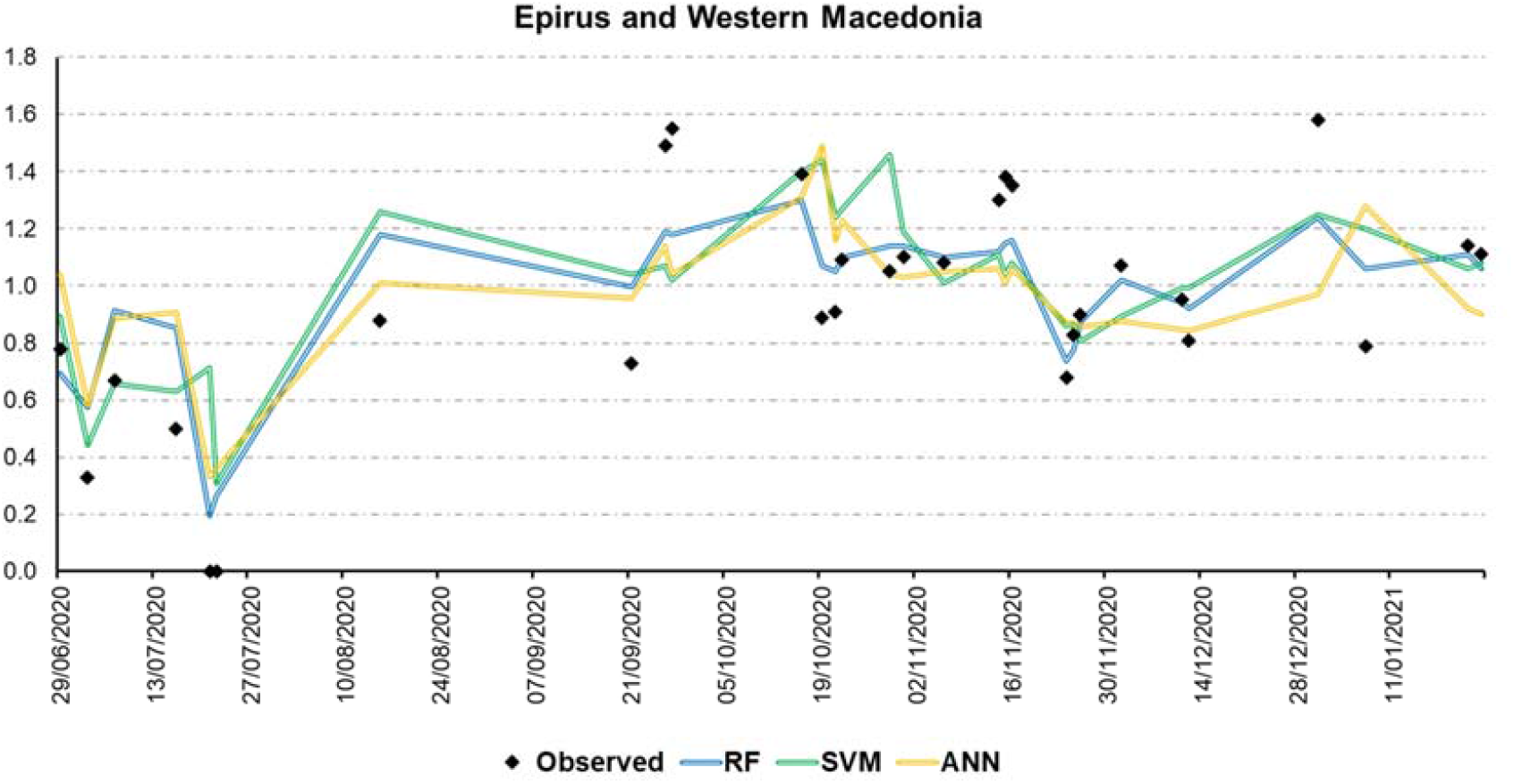
Predicted vs Observed Rt for the Epirus and Western Macedonia region.

**Figure 11.**
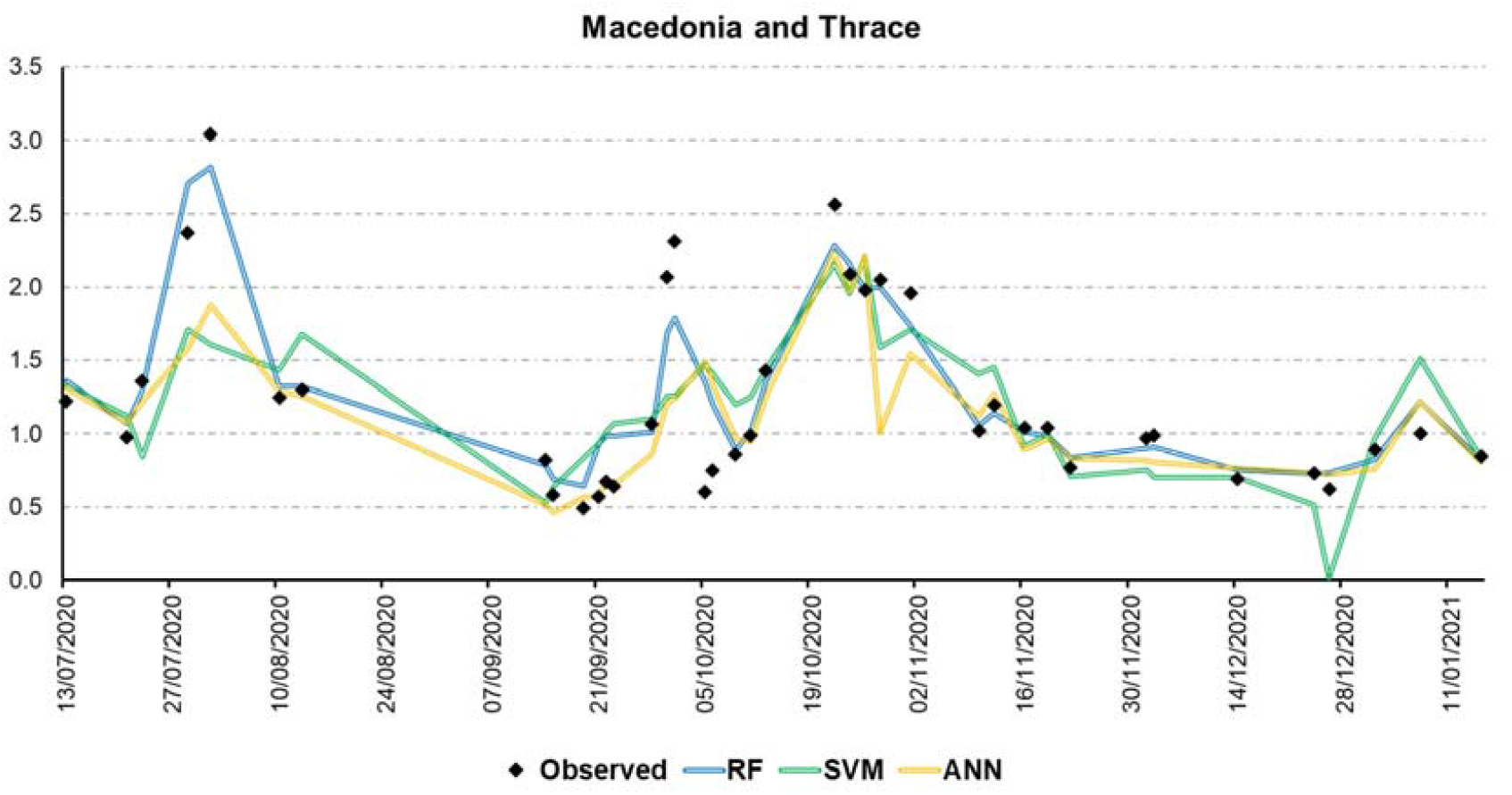
Predicted vs Observed Rt for the Macedonia and Thrace region.

**Figure 12.**
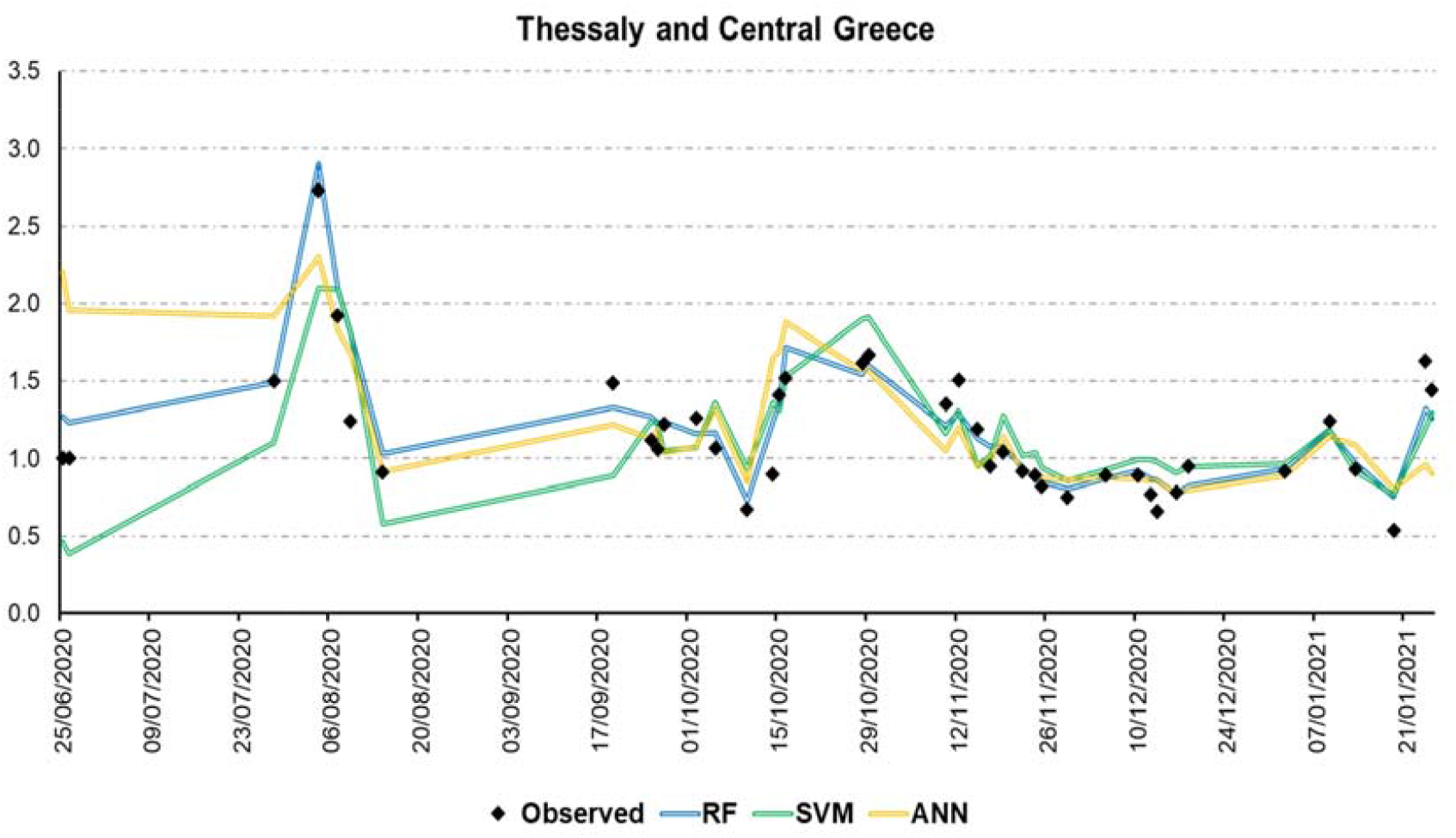
Predicted vs Observed Rt for the Thessaly and Central Greece region.

**Figure 13.**
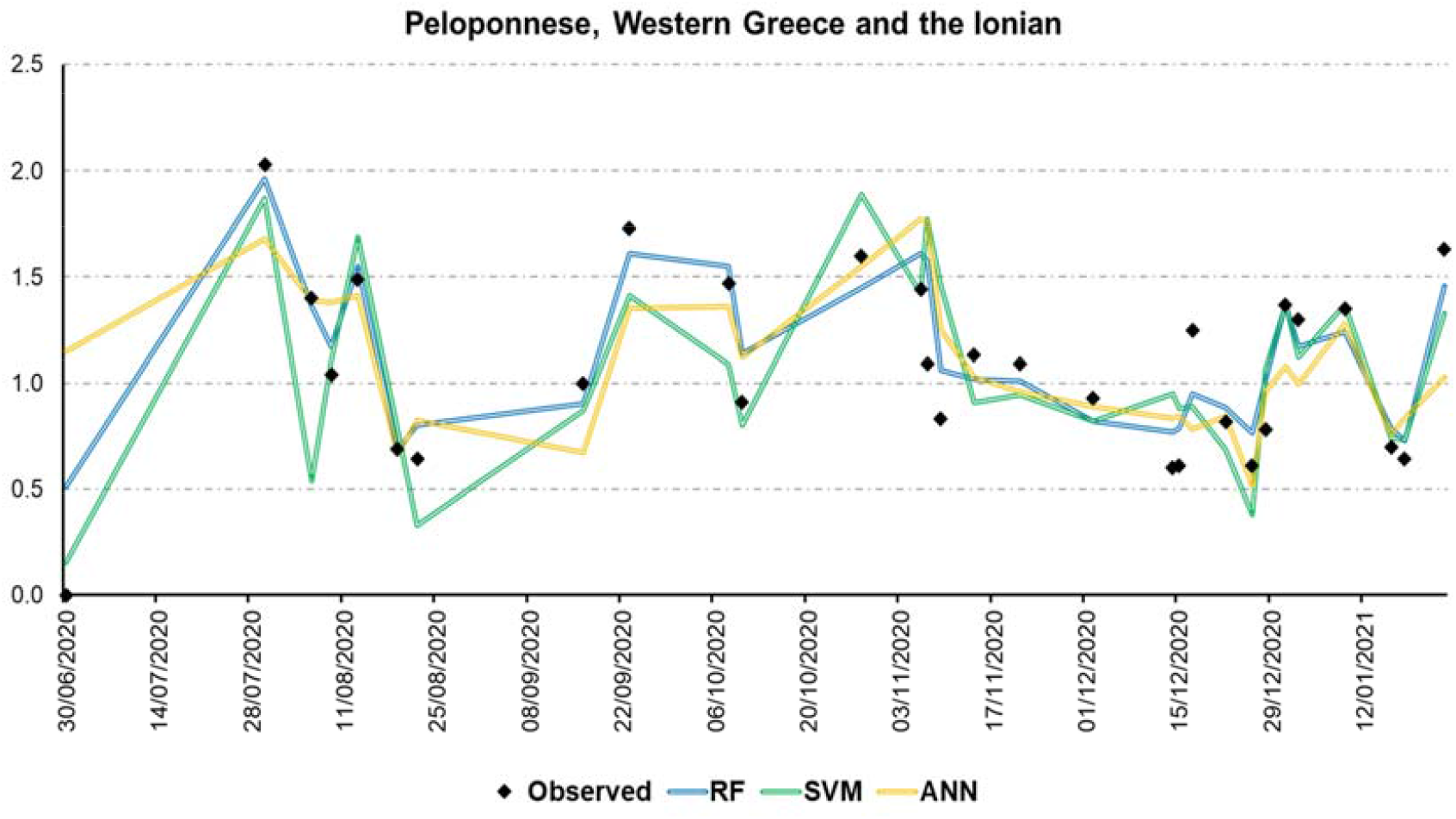
Predicted vs Observed Rt for the Peloponnese Western Greece and the Ionian region.

### 3.3 Important Attribute Analysis

The results of attribute importance analysis concerning R_t_ modelling, for the mobility categories are presented in **Figure 14**. The scores are relative, summing up to one. *Work* and *Park* categories are identified as the most important mobility features when compared to the other attributes, with values of 0.25 and 0.24, respectively. This means that when training the model, *Work* was the defining attribute in branching the RF regression trees at ∼25% of the times. According to the model built, changes in *Work* and *Park* values affect the most the predicted values of R_t_.

**Figure 14.**
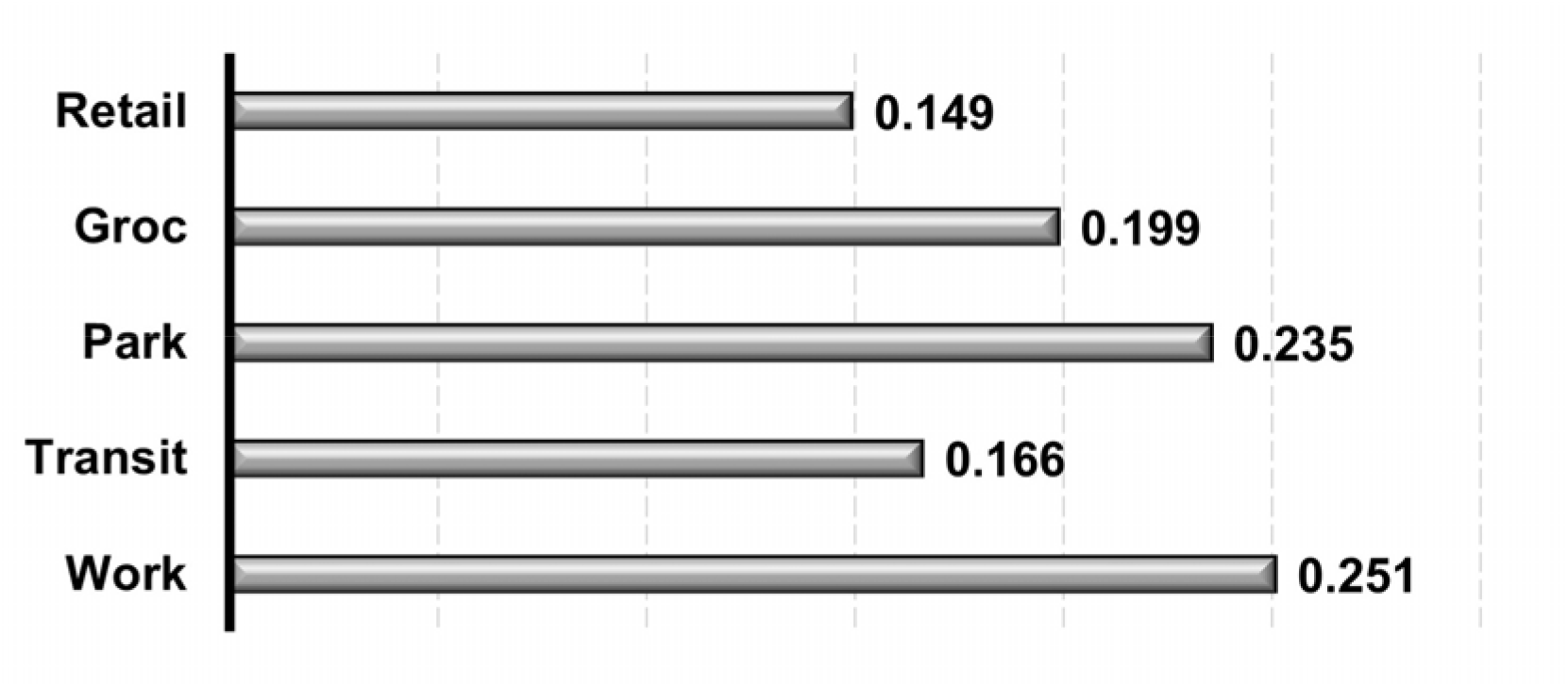
Important Attribute analysis based on RF.

## 4 Discussion

There is no universally accepted method for estimating R_t_ based on incidence data while it is much more difficult to capture its continuous temporal variation during periods of control interventions like social distancing, vaccination, changes in hygiene habits and wearing protective masks (Felizola Diniz-Filho, Jardim et al. 2020). We decided to inject the whole period data to our models without differentiating periods according to control measures implemented at the time, as we assumed that the impact of these measures compared to mobility prohibitions are minor.

We used five of the six methods for estimating R_t_ described in the EU report supporting the ECDC initiatives for COVID-19. These methods are widely used and have been implemented in a harmonized way. (Gostic, McGough et al. 2020) argue that for near real-time estimation of R_t_, the approach of Cori et al. (2013) is more suitable compared to e.g. the one of Wallinga and Teunis (2004) and also advise against using methods when their structural assumptions are not met, e.g., for Bettencourt and Ribeiro (2008). Selecting or ranking R_t_ methods was not in the scope of this study. In order to produce an estimate without having to analyse the method bias, we calculated R_t_ with several methods and used the ensemble (median value) of the results.

In order to model and subsequently forecast R_t_, we employed ML algorithms, namely RF, ANN and SVR, while we used the feature importance identification procedure based on Gini criterion, as implemented via the RF algorithm. Each one of the ML approaches that we used has its pros and cons when it comes to the modelling of datasets as the ones used in our study: RF splits each node based on the best set of predictors for each node (Breiman 2001) while bootstrap aggregation reduces the overfitting into the model. On the other hand, ANNs are highly dependent on sample size (Bataineh and Marler 2017) and small sample sizes can lead to inability for the ANN to generalize the data. Our sample size (rows) regarding the features had a proportion of 35 to 1, limiting the model ability to generalize high values of R_t_ even though still being able to approach the trend on average. Similar findings were reported in previous studies where RF showed higher robustness to regression applications compared to SVR and ANNs (Li, Yang et al. 2016, Wang, Zhou et al. 2016) due to the ability to minimize bias in multidimensional datasets (Cammarota and Pinto 2020). This outcome confirms the influence of collinearity of predictors (one predictor can predict another predictor) for SVR in a multivariate regression problem (Dormann, Elith et al. 2013) which led to higher *E* values for the margin selections. Having said that, RF was the optimal model from methodological and perspective into solving this temporal-dependent and collinear problem.

Validation metrics were chosen as MSE, MAE, R^2^, Pearson’s and Spearman’s correlation coefficient, which are the most common metrics in regression problems (Spüler, Sarasola-Sanz et al. 2015). MSE and MAE can be used to evaluate the prediction rate on average for each data point. MAE identifies the deviation from an average prediction while MSE expresses the variance of this deviation (Willmott and Matsuura 2005). Pearson’s correlation coefficient is a useful tool to evaluate the linearity between predicted values and ground truth while Spearman’s coefficient reveals the strength of monotony or the signal of it. Positive high values of Pearson’s indicate that ground truth and predicted values come into alignment. High positive values of Spearman’s CC shows that even though the model deviates from ground truth, the trendline of predictions is close (agreement in terms of the monotonic behaviour of both observations as well as model results) (Weaver, Morales et al. 2017).

Pearson’s is close to Spearman’s for all models showing that they all follow on average the temporal variation direction of R_t_ while capturing the range at a constant error. RF outperforms the other models: both Pearson and Spearman values being above 0.8, while MSE and MAE are below 0.2. ANN and SVR demonstrate a similar behaviour with the exception of a slightly higher SVM MAE and a slightly higher ANN MSE. The error metrics summarise what can be seen in **Figure 6**; RF results have the lower scatter around observed data (with a slight overestimation for lower values and a slight overestimation for higher values), while the higher SVM MAE results from the greater scatter of values around the Observed data line compared to ANN; the higher ANN MSE results from the few points with large errors (above the Observed line) even though ANN follows the Observed line more compactly.

R^2^ is moderate for SVN and ANN, a little below 0.5, but it is high, close to 0.75, for RF. This means that approximately 75% of the R_t_ observed data variance can be predicted by the RF model, suggesting that previous week changes in mobility data can explain 75% of the reproduction number daily variations. Considering other control measures independent to mobility, like social distancing, wearing masks, testing and quarantining, the mobility contribution is proven to be rather high. The result indicates that centrally imposed and monitored confinement measures are more effective than measures based on individual responsibility and involvement, at least for a population with the specific sociocultural features.

**Figure 7-Figure 13** show clearly how RF outperforms SVM and ANN. For Macedonia and Thrace, for instance, **Figure 11**, it is the only model that follows the late-July and early-October peaks. All models capture the late October increase and subsequent slow decrease, but SVM misses the Christmas holidays trend. Similar results are shown for Attica, **Figure 8**, Thessaly, **Figure 12** and Peloponnese, **Figure 13**, where RF closely approaches the R_t_ timeline, achieving the early August and after Christmas holiday peaks and even smaller peaks and plateaus. All models underperform in the cases of Crete, **Figure 9**, Aegean, **Figure 7** and Epirus and Western Macedonia, **Figure 10**, which are the most remote areas of the country, most probably due to a combination of low number of tests performed and delayed and aggregated reporting.

Coming to feature importance, *Work* and *Park* were the most impactful features for R_t_ prediction and *Retail* feature ranked as the least important one. Limitations imposed to mobility are varying in each country so direct comparisons are not reliable. For the shake of reference we note that in a similar study analysing google mobility data from 11 different countries, (Bryant and Elofsson 2020) found that grocery and pharmacy sector displayed the most significant correlations and had the highest influence in the prediction of R_0_. Other, similar studies (Wang and Yamamoto 2020, Kuo and Fu 2021) come in line with our work; predicting infection level using mobility data with or without evaluating countermeasures, such as face covering and social distancing, they demonstrated high mobility variances in *Park* ranking it as second parameter in prediction importance. It should be noted, that the least contributing mobility sector to R_t_, *Retail*, has been often the focus of control measures, sometimes limiting the number of visitors inside shops, often closing them completely. Those measures resulted in shrinking the economic activity of retail subsectors that could not compensate customer visits with online shopping, like clothing shops, for instance, that in Q4-2020 suffered losses greater than 50% in their economic activity compared to 2019^10^. Our results do not justify *Retail* to be the focus of mobility control measures to reduce R_t_, probably because face covering and social distancing had been respected inside stores more than in workplaces and parks^11^.

Training the predictive models using data from mid-June 2020 to January 2021 assumes that the dependence of the effective reproduction number to mobility does not change during this time. Besides control measures non relevant to mobility that we assumed to be minor, but we presume to account for a significant part of the 25% of the variance RF cannot capture, there are two other factors that can alter the R_t_-mobility function: herd immunity and new virus variants. Herd immunity is achieved approximately when 60-80% of population is either infected or vaccinated (Kwok et al., 2020) and it would probably compromise the model capability to predict R_t_ to the same extent. In the beginning of February 2021 only 2.79% of Greek population was vaccinated against COVID-19^12^. Although vaccination proceeds in a constant rate, a significant proportion of individuals in the general population in Greece (57.7% in a sample of 1004 respondents) are unwilling to receive a COVID-19 vaccine (Kourlaba, Kourkouni et al. 2021). According to the data used in our study around 157000 confirmed cases were reported in Greece by the end of January 2021 corresponding to ∼1.4% of the whole population. Adding vaccinated and infected numbers results in population portion insignificant to be affecting the R_t_ diagnostic and prognostic models implemented. Regarding new variants, out of the 157000 cases in late January, less than 400 were infected by the new, fast spreading UK COVID-19 variant that shows different basic reproduction numbers (40% higher), a humble 2‰ of the cases in our data that does not affect the assumption of time invariant model implementation.

## 5 Conclusions

We employed a number of Rt estimation methods based on COVID-19 test data for a period of approx. seven months (mid-June 2020 to the end of January 2021). Subsequently, we employed a ML approach for Rt modelling in order to predict the Rt value of the next day, on the basis of google mobility data of the prior week. Our results demonstrated a high ability of Rt prediction that can be used for supporting relevant short-term policy and decision making as well as scientific research related to the spread and management of the COVID-19 pandemic. To the best of our knowledge, this is the first study to predict the effective reproduction number during the COVID19 epidemic waves solely based on mobility information and for subnational regions of Greece. The results differentiate between activities and places guiding authorities to be precise and effective on their future public health interventions, both at the national and regional level. The predictive model assumes low herd immunity and a constant dominant variant; variations from these assumptions would probably compromise the model capability to predict R_t_ to the same extent, unless such information is made available for the same spatial and temporal resolution used in our study.

## Data Availability

Data available from the authors upon requrest

## Acknowledgments

The authors would like to thank Drs T. Asikainen and A. Annunziato from the European Commission Joint Research Centre, for their assistance modifying and implementing the software of R_t_ calculation from confirmed cases data.

## Footnotes

1 *https://www.worldometers.info/coronavirus/country/greece/*

2 *https://graphics.reuters.com/world-coronavirus-tracker-and-maps/countries-and-territories/greece/*

3 *https://github.com/iMEdD-Lab/open-data/tree/master/COVID-19*

4 *https://ourworldindata.org/coronavirus-testing*

5 *https://ww.scienzainrete.it/articolo/cruscotto-monitorare-levoluzione-dellepidemia/cesare-cislaghi/2020-04-09*

6 *https://www.rki.de/DE/Content/Infekt/EpidBull/Archiv/2020/Ausgaben/17_20.html*

7 *https://github.com/ec-jrc/COVID-19/tree/master/programs/ReprNumber*

8 *https://www.latinamerica.undp.org/content/rblac/en/home/presscenter/director-s-graph-for-thought/home-alone---sustaining-compliance-with-prolonged-covid-19-stay-.html*

9 *In the context of model validation “observed” refers to the R*_*t*_ *values calculated by the five standard methods described in 2.2 and 3.1 based on numbers of confirmed cases*.

11 https://www.capital.gr/oikonomia/3527629/elstat-meiothike-8-6-o-kuklos-ergasion-sto-lianiko-emporio-to-2020.

12 *https://ourworldindata.org/covid-vaccinations*

